# A Multicenter Study of Coronavirus Disease 2019 Outcomes of Cancer Patients in Wuhan, China

**DOI:** 10.1101/2020.03.21.20037127

**Authors:** Hongyan Zhang, Linwei Wang, Yuanyuan Chen, Xiaokun Shen, Qun Wang, Youqin Yan, Yi Yu, Qiuji Wu, Yahua Zhong, Melvin L.K. Chua, Conghua Xie

**Author notes:** **Corresponding Authors:** Prof. Conghua Xie, Department of Radiation and Medical Oncology, Zhongnan Hospital of Wuhan University, No. 169, Donghu Road, Wuchang District, Wuhan, Hubei Province, 430071, China. Dr Melvin L.K. Chua, Division of Radiation Oncology, National Cancer Center Singapore, 11 Hospital Dr, Singapore 169610. Prof. Yahua Zhong, Department of Radiation and Medical Oncology, Zhongnan Hospital of Wuhan University, No. 169, Donghu Road, Wuchang District, Wuhan, Hubei Province, 430071, China. Co-first authors (Hongyan Zhang, Linwei Wang, Yuanyuan Chen, Xiaokun Shen). Co-senior authors (Yahua Zhong, Melvin L.K. Chua, Conghua Xie).

## Abstract

**Background:** At present, there is a global pandemic of coronavirus disease 2019 (COVID-19) pneumonia. For COVID-19 patients, cancer is a coexisting disease that should not be Here, we conducted a multicenter retrospective study to show the clinical information and outcomes of cancer patients infected with COVID-19.

**Measurements:** Medical records of COVID-19 patients with cancer admitted to hospitals from Jan 5, 2020 to Feb 18, 2020 were collected.

**Results:** Of the 67 patients (median age: 66 years), the median age of patients with severe illness was older than that of patients with mild symptoms (*P*<0.001). The proportion of severe patients had co-morbidities was higher than patients with mild disease (*P*=0.004). During the treatment of COVID-19 pneumonia, tumor progression and recurrence was not observed for those patients still at the anticancer treatment phase. Lymphocytopenia was the main laboratory finding accompanying increased C-reactive protein and procalcitonin in cancer patients, especially in severe cases. By Mar 10, 2020, 18 (26.9%) patients died from COVID-19. The median age of survivors was younger than that of deaths (*P*=0.014). Lung cancer (n=15, 22.4%) was the most common cancer type and accounted for the highest proportion COVID-19 resulted deaths (33.3%, 5/15). We observed a tendency that patients at the follow-up phase had a better prognosis than that at anticancer treatment phase (*P*=0.095).

**Conclusion:** This study showed COVID-19 patients with cancer seem to have a higher proportion of severe cases and poorer prognosis. We should pay more intensive attentions to cancer patients infected with COVID-19.

## Introduction

An outbreak of coronavirus disease 2019 (COVID-19) has occurred in China and the rest of world since Dec 2019, which was linked to the severe adult respiratory syndrome coronavirus 2 (SARS-CoV-2).^1-4^. According to the situation report from World Health Organization (WHO), COVID-19 has caused 332930 confirmed cases and 14510 deaths in global by Mar 23, 2020.^5^ WHO has declared COVID-19 a Public Health Emergency of International Concern, and raised its risk assessment from “high” to “very high”. And the COVID-19 has been becoming a global pandemic.

Considering that the number of patients infected by SARS-CoV-2 is large and still increasing, cancer is a coexisting disease that should not be neglected. Many patients with cancer are at the risk of SARS-CoV-2 infection. Previous study including 18 cancer patients found that patients with cancer had a higher risk of COVID-19 and deteriorated more rapidly than individuals without cancer in 1099 cases.^6,7^ There were 107 patients with malignancy among a total of 72314 patient records diagnosed as of February 11, 2020.^8^ However, we still could not fully understand the clinical characteristics of cancer patients with COVID-19 from the above-mentioned study due to limited sample size. Here, we conducted a multicenter retrospective study to show the clinical information and outcomes of 67 cancer patients infected with COVID-19 in 1548 cases from four designated COVID-19 hospitals.

## Methods

### Study design and participants

For this multicenter, retrospective, observational study, we identified 67 cancer patients with COVID-19 from 1548 inpatients in four designated hospitals (Zhongnan Hospital of Wuhan University, the Fifth Hospital of Wuhan, the Seventh Hospital of Wuhan, and Wuhan Hankou Hospital). Patients were admitted to hospital from Jan 5, 2020 to Feb 18, 2020. This study was approved by the ethics commissions of Zhongnan Hospital of Wuhan University, with a waiver of informed consent due to a public health outbreak investigation.

### Definitions

COVID-19 were diagnosed by real-time reverse transcription polymerase chain reaction assay for SARS-CoV-2 according to recommended protocol.^9^ A small number of cases were described as clinically confirmed cases based on Diagnosis and Treatment Program of 2019 New Coronavirus Pneumonia (v5.0 Feb 8, 2020) by National Health and Health Commission of China.^10^ We also categorized the patients according to the disease severity. A mild case was defined as a confirmed case with common symptoms and mild radiographic evidence of pneumonia, while a severe case with dyspnea or respiratory failure. All cancer patients were divided into follow-up group and ongoing treatment group. Follow-up group was defined as routinely radical treatments were finished more than one month, and ongoing treatment group was defined as anticancer therapy is required, not finished or finished in one month.

### Data collection

Detailed information of medical records, including epidemiological, demographics, exposure history, smoking history, chronic medical illness, signs and symptoms on admission, comorbidity, chest radiography and CT findings, laboratory findings and treatment measures were collected in this study. Clinical outcomes were followed up to Mar 10, 2020. All data was checked by two researchers.

### Statistical analysis

We compared the characteristics between severe and mild cases, survivors and non-survivors. Continuous variables were described using mean (standard deviation, SD) if they are normally distributed, median (range) if they are not. We expressed descriptive data as count (%) for categorical variables. All statistical analyses were performed using IBM SPSS statistics (version 23.0 for Windows). Two-sided *P* value less than 0.05 was considered statistically significant.

## Results

### Clinical characteristics of cancer patients with COVID-19 pneumonia

By Feb 18, 2020, we identified 67 cancer patients with COVID-19 from four designated hospital (Zhongnan Hospital of Wuhan University, the Fifth Hospital of Wuhan, the Seventh Hospital of Wuhan, Wuhan Hankou Hospital). Among them, 58 (86.6%) patients had a positive real-time reverse transcription polymerase chain reaction (RT-PCR) assay for SARS-CoV-2, and 9 (13.4%) patients were clinical diagnosed cases. 94.0% (64/67) of patients had radiological changes on chest CT scan. Hospital-acquired infection occurred in 9 (13.4%) patients. Detailed clinical information about these patients was summarised in **Supplementary file 1**.

Clinical characteristics of the 67 COVID-19 cases are summarised in **Table 1**. Of the 67 cancer patients involved in our study, 41 (61.2%) patients were male. The median age of patients was 66 years old (range: 37-90). In terms of disease severity, 35 (52.2%) patients had a mild illness, while 32 (47.8%) patients had a severe illness. The median age of patients who had severe illness was older than that of patients who had mild symptoms (69 vs. 64 years, *P*<0.001). Additionally, we observed that a higher proportion of severe patients had co-morbidities compared to patients with mild disease (71.9% [severe] vs 37.1% [mild], *P*=0.004). Of note, three of the severe cases had concurrent chronic obstructive pulmonary disease (COPD), and these individuals did not survive from the COVID-19 pneumonia. Nine (13.4%) patients have a smoking history. Forty-three (64.2%) patients had other concurrent chronic diseases, of which hypertension (53.7%, n=36), diabetes (19.4%, n=13), and coronary heart disease (11.9%, n=8) were the most common. In addition, 23 (23/36, 66.7%) patients with concurrent hypertension were severe, accounting for 71.9% (23/32) of all the severe patients in our cohort.

### Association of cancer diagnosis with COVID-19 pneumonia

Among the cancer diagnosed cancers, lung cancer (n=15, 22.4%) was the most common cancer type. Forty-four (65.7%) patients were cancer survivors on routine follow-up after radical treatment. Twenty-three (34.3%) patient had received anticancer treatment recently (**Table 1**). Only two patients were still ongoing anticancer treatment after the SARS-CoV-2 infection. Among those 23 patients, 9 cases were receiving chemotherapy-based comprehensive therapies, 7 cases were during perioperative period, 3 cases were receiving supportive treatment, 2 cases were receiving radiotherapy combined with targeted therapy or with endocrinotherapy, and the rest 2 patients were receiving endocrinotherapy or immunotherapy. During the treatment of COVID-19 pneumonia, no tumor progression or recurrence was observed in all the patients.

### Clinical progression and treatment

About 70% of these patients had fever (n=53, 79.1%) and cough (n=50, 74.6%) (**Table 2**). More than half of them had shortness of breath (n=44, 65.7%). Other clinical symptoms included fatigue (n=30, 44.8%), sputum production (n=30, 44.8%), dyspnea (n=22, 32.8%), diarrhea (n=11, 16.4%), nausea and vomiting (n=10, 14.9%), myalgia (n=6, 9.0%) and hemoptysis (n=3, 4.5%). Severe patients and non-survivors had a higher proportion of dyspnea at admission (mild vs. severe, 11.4% vs. 56.3%; survivors vs. non-survivors, 20.4% vs. 66.7%, *P*<0.05). The other baseline symptoms were similar between mild and severe cases in general. According to the results of computed tomography (CT) scan, 27 (42.2%) patients had severe baseline CT manifestations. **Table 3** showed the baseline blood test results. Similar to general infected patients, lymphocytopenia was the main laboratory finding accompanied by increased C-reactive protein and procalcitonin in cancer patients, especially in severe cases. In addition, we observed decreased platelets and increased blood urea nitrogen and lactate dehydrogenase in severe patients/non-survivors.

The landscape of treatment was displayed in **Table 2**. Fifty-seven (85.1%) patients received antiviral treatment. Thirty (44.8%) patients were also treated with glucocorticoids, and 14 (20.9%) patients treated with intravenous immunoglobulin therapy. More than 70% (n=49, 73.1%) of patients received oxygen therapy. Twenty (29.9%) patients used non-invasive ventilator mechanical ventilation and 8 (11.9%) patients used an invasive ventilator to assist ventilation.

#### Outcomes of cancer COVID-19 patients

By Mar 10, 2020, 14 (20.9%) patients had developed acute respiratory distress syndrome (ARDS). The other common serious complication was heart failure (n=11, 16.4%). Two (3.0%) patients presented with acute renal injury. In our cohort, 18 (26.9%) patients died from COVID-19, and 39 (58.2%) patients have been discharged. The remaining 10 (14.9%) patients were still in hospital for treatment. The case-fatality rate (CRF) in this study for overall patients, patients at anticancer treatment phase, and at the follow-up phase is 26.9% (18/67), 39.1% (9/23) and 20.5% (9/44), respectively. We observed a tendency that patients at the follow-up phase might have a better prognosis than under treatment, but there was statistically significance (**Figure 1**, *P*=0.095). The median time from onset to severe events for severe cases was 10 (range: 1-21) days. The median durations from onset to recovery and death were 31 (range: 8-53) and 20 (range: 6-45) days, respectively. The median age of survivors was younger than that of deaths (65 [37-83] vs 69.5 [52-98], *P*=0.014).

**Figure 1.**
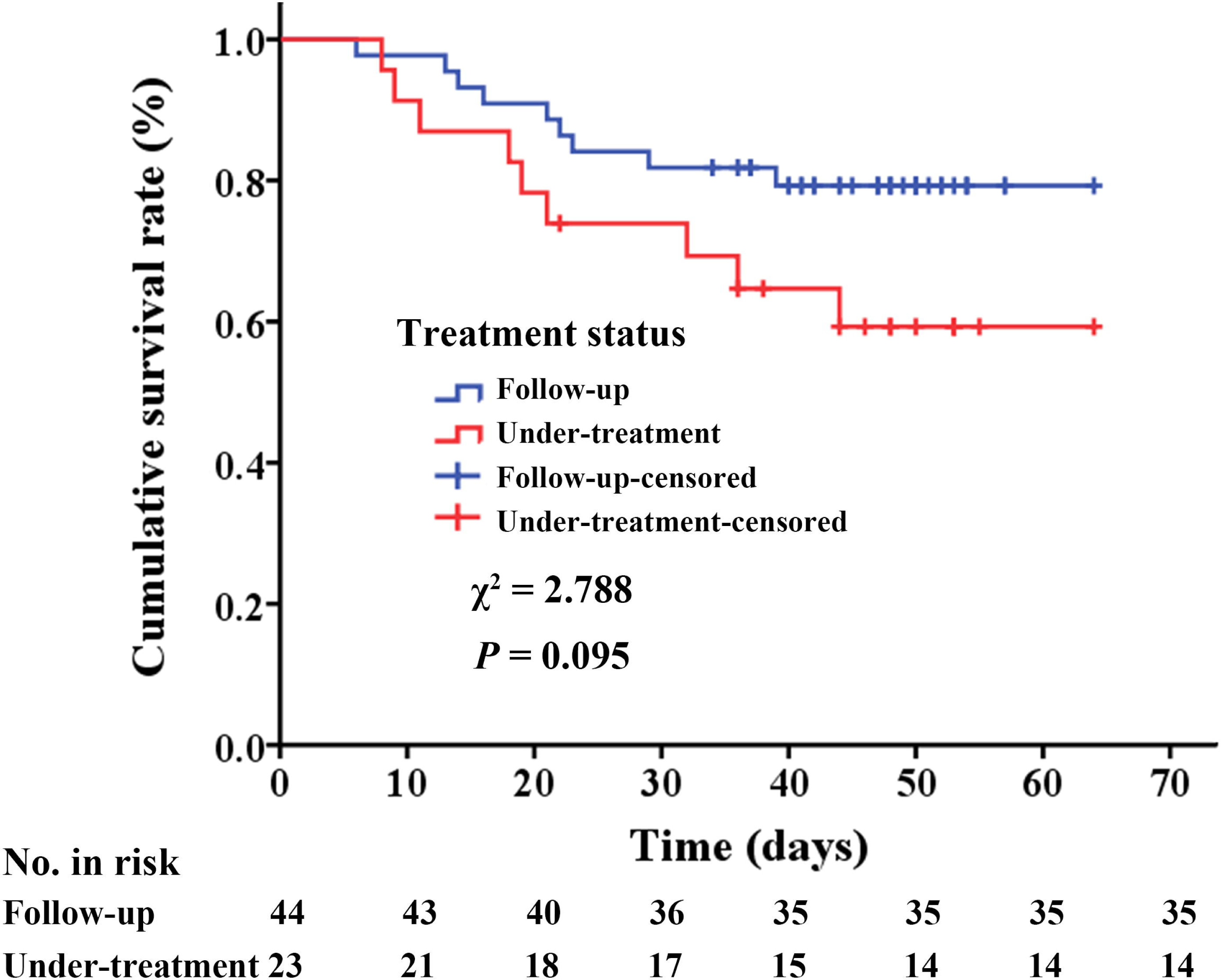
Survival analysis of cancer patients with COVID-19 at anticancer treatment phase and follow-up phase.

### Comparison of different cancer types

To clearly compare clinical characteristics of lung cancer with other cancers, distributions of cancers for different treatment phases, disease severity, age group and disease phase were showed in **Figure 2**. Number of COVID-19 patients with lung cancer under treatment is twice of patients in follow-up (**Figure 2A**). Digestive system cancer patients ranked the highest proportion of severe condition (**Figure 2B**). Proportions of lung (12/15, 80%) and digestive system cancers (14/17, 82.4%) with age ≥ 65 were higher than of other cancers (**Figure 2C**). Up to the follow-up deadline, the CFR of COVID-19 patients with lung cancer (5/15, 33.3%) was higher than that of digestive system cancers (5/17, 29.4%) and other cancers (22.9%) (**Figure 2D**). Of note, all deaths (5 cases) in patients with lung cancer were at under treatment phase.

**Figure 2.**
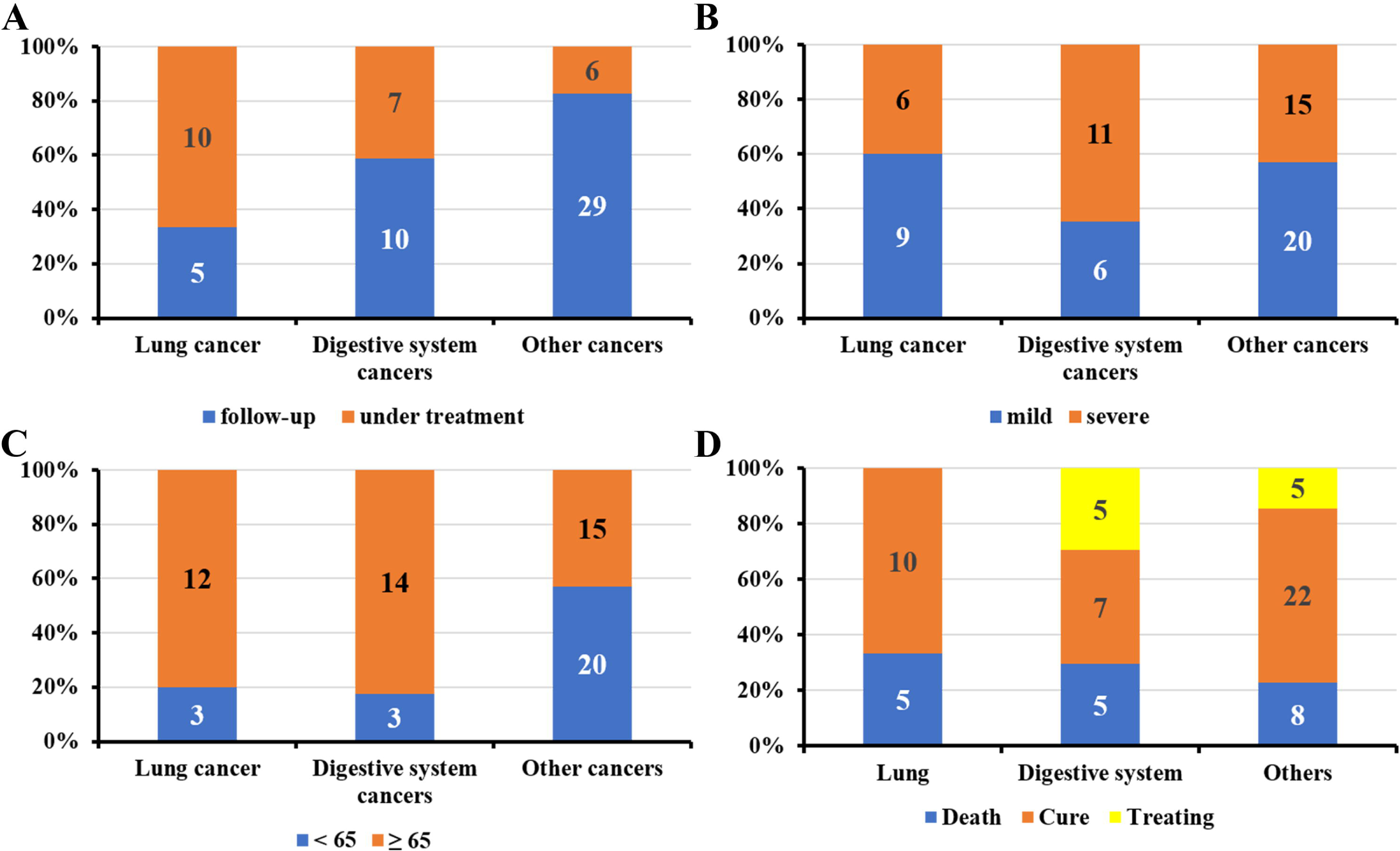
Comparisons among lung, digestive system and other type cancers. Distribution of different cancers for tumor phase (Panel A), disease severity (Panel B), age group (Panel C) and disease phase (Panel D)

## Discussion

In this multicenter retrospective study, we reported the largest cohort of cancer patients with COVID-19 to our knowledge. Up to Mar 10, 18 (26.9%) patients died from COVID-19, 39 (58.2%) patients have been discharged, and 10 (14.9%) patients were still in hospital for treatment in our cohort. Perhaps most notably, we found that about half of the cancer patients with COVID-19 developed severe illness, which is higher than the proportion in general population (15.6%, 245/1572 in Guan et.al,^11^ **Supplementary Table S1**). According to a nationwide study based on data regarding 1099 patients with COVID-19 in China, about 3.4% of COVID-19 patients develop ARDS,^7^ while the incidence rate of ARDS in our cancer cohort was 16.42%. The CFR rate of cancer patients infected by SARS-CoV-2 was higher than that of the overall population. According to the situation report from the WHO, 153517 cases of COVID-19 had been confirmed all around the world up to Mar 15, 2020, with a CFR rate of approximately 3.7%.^5^ In contrast, the crude CFR in our cohort was about 26.9%. Our findings provided evidence that cancer patients with COVID-19 could have a poorer outcome, which was consistent with the previous study.^6^

In this study, the CFR of patients receiving ongoing anticancer treatment recently (39.1%, 9/23) was higher than those who were merely on follow-up post-cancer treatment in the routine follow-up phase after radical treatment (20.5%, 9/44). Furthermore, we observed a tendency that patients at the follow-up phase may have a better prognosis than those under treatment (**Figure 1**, *P*=0.095). Those results provide further evidence to demonstrate that patients receiving ongoing anticancer treatment recently have a poorer prognosis than those patients at follow-up phase. It may a result of immune compromising effect led by anticancer treatments.^12-14^ Noteworthy, this study showed that lung cancer was the most common cancer type (accounted for 22.4%), which may have higher susceptibility SARS-CoV-2 infection than other type cancers. There may be two main reasons: (1) lung cancer accounts for a large proportion of all cancers; (2) 10 cases lung cancer patients were ongoing anticancer treatment, and they may have high risks of nosocomial infection and low immunity to SARS-CoV viral particles. Meanwhile, all 5 deaths with lung cancer is at anticancer treatment phase. This indicates that patients with lung cancer at anticancer treatment phase might have poor prognosis. The proportion of severe cases among different kinds of cancer types ranged from 25.0% to 64.7%, of which digestive system cancer patients ranked the highest. However, it is still unclear whether there are any differences in the severity of SARS-CoV-2 infection among other types of cancer. More data is needed to further verify our findings.

As reported in previous studies,^4,15,16^ male and older patients had a higher risk of SARS-CoV-2 infection, which was validated in our cancer cohort. Patients in our cancer cohort were older overall than patients in previous reports.^4,15,16^ It may because those patients have accompanying cancer. In addition, we found that severe patients tended to be older than patients with mild illness, which indicated that older patients were at an increased risk of being critically ill. Patients who had other chronic illnesses were also at a higher risk of being severe according to previous studies.^4,15,17^ In our cohort, we also observed that a higher proportion of severe patients had co-morbidities compared to patients with mild disease (*P*=0.004). Particularly, about 70% of patients with concurrent hypertension were severe, accounting for the majority of the severe patients in our study. Recent studies suggested that angiotensin-converting enzyme-2 (ACE2) could be the host receptor for SARS-CoV-2 with high binding affinity.^18,19^ Besides, the expression of ACE2 in tumor tissues could be higher than adjacent tissues and there is expression heterogeneity across different cancer types.^20^

In the clinical courses of cancer patients with COVID-19, lymphocytopenia was observed, especially in severe patients/non-survivors, which might result from targeted damages by SARS-CoV viral particles (21). Our findings were consistent with previous studies on COVID-19,^4,15^ indicating that lymphocytopenia reflects the severity of COVID-19 in a way. As for treatment, antiviral regimens were used in most patients in our study. To our knowledge, there was no specific remedy for COVID-19 yet.^7,22^ Therefore, supportive treatment has been adopted as the mainstay of therapy. In our cohort, about 73.1% of cancer patients with COVID-19 received oxygen therapy. About 40% of the patients received mechanical ventilation.

Hospital-acquired transmission accounted for 13.4% of our cancer patients. This data was consistent with the previous report of 138 hospitalized patients from one of our center, in which 12.3% of in-hospital patients were infected with SARS-CoV-2 through hospital-acquired transmission.^17^ It indicated that hospital-acquired transmission was also an important way of infection in cancer patients. In addition, cancer patients are at a higher risk of COVID-19, even if they have completed the anticancer treatment. More than 60% (65.6%) of cancer patients in our study were at the follow-up phase after radical anticancer treatment. For regular anticancer treatment and follow up, cancer patients need to visit hospital frequently, which increases the risk of SARS-CoV-2 exposure and infection. Besides, cancer patients with COVID-19 might have more complications due to their old age, weakened immune system and the side effects of undergoing treatments.^12,14^ Therefore, we recommend systemic screening should be conducted among hospitalized cancer patients. In our study, we also observed that ongoing anticancer treatment was interrupted in most patients because of the diagnosis of COVID-19. Fortunately, we had not observed tumor progression during the treatment of COVID-19 pneumonia. On one hand, appropriately delaying treatment and reductive frequency of visit hospital for cancer patients is feasible. On the other hand, medical oncologists are recommended to provide professional consultation for COVID-19 cancer patients to ensure essential anticancer treatments.

This study has several limitations. First, this is a retrospective study with only 67 cases from four hospitals. Although it is the largest cancer patient cohort of COVID-19 up to now. The results may not be representative of the entire cancer patient population and should be interpreted with caution. Second, some specific clinical information was insufficient and whether COVID-19 had further influence on the therapy and prognosis for cancer patients was not analysed in this study. Therefore, an extended follow-up should be recommended for these patients. In the future, a larger COVID-19 cancer patient cohort from multicenter is needed to explore more detail information of cancer patients.

## Conclusion

In this study, we reported the clinical characteristics and outcomes of COVID-19 in cancer patients. Both patients within the follow-up phase and ongoing anticancer treatment phase were at the risk of COVID-19. This study showed COVID-19 patients with cancer seem to have a higher proportion of severe cases and poorer prognosis. The tendency of poor prognosis was more obvious in patients at anticancer treatment phase. We should pay more intensive attention to cancer patients, especially those with common risk factors of COVID-19.

## Data Availability

The data of this study is available to all authorized members.

## Contributors

HYZ, LWW, YYC, QW, YQY and YY had full access to all the data in the study and takes responsibility for the integrity of the data and the accuracy of the data analysis.

Concept and design: HYZ, XKS, MLC, YHZ, CHX.

Acquisition, analysis, or interpretation of data: HYZ, LWW, YYC.

Drafting of the manuscript: HYZ, LWW, XKS.

Critical revision of the manuscript for important intellectual content: YHZ, MLK, CHX.

Statistical analysis: QJW.

Obtained funding: CHX.

Administrative, technical, or material support: YHZ, CHX.

Supervision: YHZ, MLK, CHX.

## Reproducible Research Statement

Study protocol: Not applicable.

## Acknowledgments

We thank all patients and frontline healthcare workers involved in the study.

## Primary Funding Source

This study was funded by Health Commission of Hubei Province Scientific Research Project (WJ2019H002) and Health Commission of Hubei Province Medical Leading Talent Project.

## Ethical statement

Approval of the study protocol was obtained from the Institutional Ethics Committee of hospitals (No. 2020041). And informed consent of this study was waived. The study was undertaken according to the ethical standards of the World Medical Association Declaration of Helsinki.

